# Leveraging Health Systems Data to Characterize a Large Effect Variant Conferring Risk for Liver Disease in Puerto Ricans

**DOI:** 10.1101/2021.03.31.21254662

**Authors:** Gillian M. Belbin, Stephanie Rutledge, Tetyana Dodatko, Sinead Cullina, Michael C. Turchin, Sumita Kohli, Denis Torre, Muh-Ching Yee, Christopher R. Gignoux, Noura S. Abul-Husn, Sander M. Houten, Eimear E. Kenny

## Abstract

Broad-scale adoption of genomic data in health systems offers opportunities for extending methods for the discovery of variation linked to underlying genomic disease risk. We applied a population-scale linkage mapping approach in a large multi-ethnic biobank to a spectrum of disease outcomes derived from Electronic Health Records (EHRs) and uncovered a risk locus for liver disease. We used genome sequencing and *in silico* approaches to fine-map the signal to a non-coding variant (*c*.*2784-12T*>*C)* in the gene *ABCB4. In vitro* analysis confirmed the variant disrupted splicing of the ABCB4 pre-mRNA. Four of five homozygotes had evidence of advanced liver disease, and there was a significant association with liver disease among heterozygotes, suggesting the variant is linked to increased risk of liver disease in an allele dose-dependent manner. Population-level screening revealed the variant to be at a carrier rate of 1.95% in Puerto Rican individuals, likely as the result of a Puerto Rican founder effect. This work demonstrates that integrating EHR and genomic data at a population-scale can facilitate novel strategies for understanding the continuum of genomic risk for common diseases, particularly in populations underrepresented in genomic medicine.

## Introduction

Genetic identification of monogenic disease historically relied on tracking the co-segregation of genomic segments and disease-state through familial pedigrees, in a process known as linkage mapping [1,2]. This approach is typically followed by localized sequencing to reveal the disease causing variant and confirmatory functional studies *in vitro* or in animal models. This strategy has been used successfully throughout the late 20th century to uncover thousands of loci underlying suspected, rare genetic disorders[3]. More recently, next generation sequencing technologies have led to the identification of the genetic etiology of disease through the direct sequencing of patient exomes and genomes in close pedigree structures[4]. Genomic technologies have also been applied in health systems to uncover unknown pathogenic variants and streamline diagnosis[5] and to refine our understanding of the penetrance and frequency of pathogenic variants at a population-level[6]. However, the preponderance of genome sequencing and genomic medicine research have been performed in populations of European descent, and there is a lag in genomic sequence data available for, and studies directed at, understanding monogenic disorders in non-European populations[7].

The growth of large-scale biobanks linked to health systems data in recent years has opened new avenues to uncovering the etiology of known and novel monogenic disorders[8]. With some exceptions[9][10], the majority of genomic data generated in biobanks worldwide is on low-cost genotype arrays rather than genome sequencing and many biobanks are designed for population-based recruitment rather than being disease or pedigree focused. However, by leveraging array data in population-based biobanks, it is possible to calculate haplotypes of the genome that have been co-inherited from a recent common ancestor Identical-by-Descent (IBD)[11]. Using this strategy, genealogical relationships can be captured locally along the genome among distantly, or putatively unrelated members of a population, which are particularly enriched in founder populations[12–14][15]. IBD-haplotypes have the potential to harbour rare alleles that are not directly ascertained on genotyping arrays, facilitating association mapping of rare variants even when they are not directly observed[12], or are too rare or population-private to be readily imputable with currently existing reference panels; this approach is known as population-scale linkage or IBD-mapping[15,16]. This property of IBD makes it especially useful for rare variant based associations in diverse and understudied founder populations, for which deep genome sequencing datasets may not be available. Furthemore, previous studies have leveraged HER data in concert with genomic data to demonstrate the ubiquity and potential under-recognition of monogenic forms of disease in patient populations[6,17]. We previously applied IBD-mapping to height in a Puerto Rican (PR) founder population in New York City and identified a monogenic variant underlying the skeletal disorder Steel Syndrome[18], demonstrating the power for discovery of monogenic variants underlying monogenic disorders.

Here we expand our previous approach by systematically associating IBD haplotypes with the full spectrum of EHR derived phenotypes in the large founder population of PR and PR descent participants in the diverse, multi-ethnic Bio*Me* biobank in New York City. We performed a Phenome-Wide Association Study (PheWAS)[19] of IBD-haplotypes under a recessive model in the PR founder population and identified a significant association between homologous IBD sharing at the locus 7q21.12 and severe liver disease. Fine-mapping of the IBD-haplotypic region uncovered a rare variant in *ABCB4* (*ABCB4:c*.*2784-12T*>*C*; rs201498350), a gene known to play a causal role in multiple forms of hepatobiliary disease[20]. *In vitro* analysis demonstrated that this variant disrupted splicing, leading to an ABCB4 protein product lacking exon 23. Manual chart review of these patients revealed evidence of severe liver diseases in four of five homozygotes. We also investigated the impact of harboring one copy of *c*.*2784-12T*>*C* via a combination of PheWAS, analysis of liver function tests, and manual chart review, revealing an increased risk of liver disease in heterozygotes. Furthermore, population-level screening revealed the variant to be common in PR (carrier rate of ∼1.9%), while rare (<1%) in other global populations. These analyses provide a methodological framework for bridging statistical genetics and clinical genomics, and demonstrate that EHR-embedded, population-level research can elucidate the continuum of genomic risk for liver disease.

## Results

### Inference of Identity-by-Descent Haplotypes in PR population

We previously inferred Identity-by-Descent (IBD) sharing across Bio*Me* and used IBD haplotypes to cluster patients into communities linked by recent shared ancestry, as described in Belbin et al. 2019[21]. By using this method, we identified a community of individuals of Puerto Rican (PR) ancestry, and observed elevated IBD sharing within this group, suggestive of a founder effect. We clustered IBD haplotypes locally along the genome by homology and identified 4526956 homologous IBD-clusters within the PR population. Examining the frequency spectrum of these haplotypic alleles, we observed most to be rare (median haplotypic frequency=0.04%, **Supplementary Figure 1**). We hypothesized that we may be able to leverage these haplotypic alleles as proxies for unobserved rare variants in an association testing framework designed for discovery of monogenic recessive disorders (**Figure 1**).

**Figure 1.**
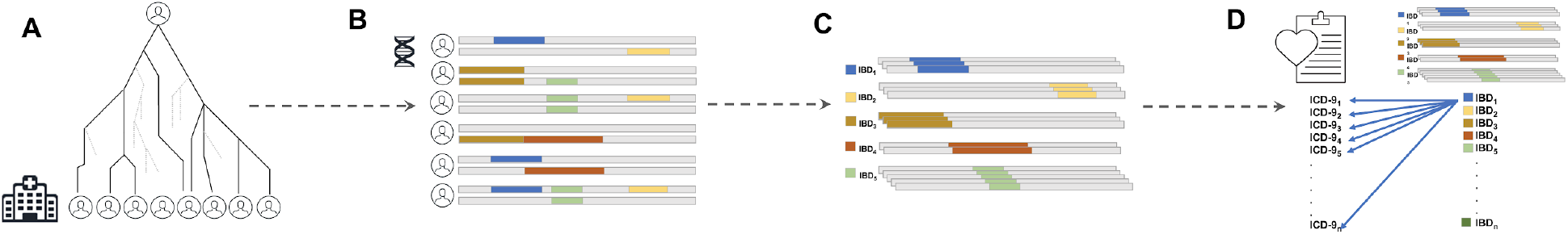
Framework for population scale linkage analysis in a health system. **A)** Distant, cryptic genealogical relationships are present in large, putatively unrelated patient populations. **B)** This leads to the presence of genomic patterns of Identity-by-Descent (IBD) haplotype sharing across distantly related patients. These haplotypes have been co-inherited from a recent, shared common ancestor at some point in recent genealogical history. IBD can be inferred from phased genotype data and clustered into homologous groups. **C)** Different coloured IBD-haplotypes represent different groups of IBD that have each been co-inherited identically by multiple individuals and that can be clustered together according to homology. **D)** Population-scale linkage mapping performed via Phenome-Wide Association (PheWAS) by extracting phenotypes from the Electronic Health Records (EHR) in the form of ICD-9 billing codes, and systematically testing for association between all IBD-haplotype clusters and all ICD-9 code outcomes.

### Phenome Wide Association of Identity-by-Descent Haplotypes in Puerto Rican Community

To systematically explore the relationship between haplotypic alleles and EHR derived health outcomes, we performed a PheWAS under a recessive model implementing the Saddle Point Approximation, which accommodates for rare observations and instances of extreme case-control imbalance. Because our method depends on leveraging cryptic relatedness, we applied our approach specifically within PR Bio*Me* participants, on the basis of previous observations of a founder effect within this group [18]. In our model, the haplotypic alleles served as the primary predictor variable and ICD-9 billing codes served as the outcome variable. We restricted analysis to 754 haplotypic alleles for which there were at least 3 observations of individuals that were homozygous for a shared IBD haplotype, and systematically tested these for association against each ICD-9 code (n=3,679,520 tests in total). Only one association achieved study-wide significance (SWS, threshold: p<1.4×10^−8^), an association at a haplotypic allele at 7q21.12 (p<2.9×10^−9^, haplotypic frequency=0.7%) (**Figure 2A**). The significant haplotypic allele represented 3 individuals who were each homozygous for a homologous segment of IBD at the region, and all of whom had EHR record of the rare ICD-9 code “571.6” (which encodes for “Biliary Cirrhosis”). While not study-wide significant, the haplotypic allele was also associated with the ICD-9 code “576.1” (which encodes for ‘cholangitis’; p <9.9×10^−8^). In addition to the three individuals who were homozygous for the IBD haplotype at 7q21.12, N=70 individuals carried the haplotype in the heterozygous state. The significant haplotypic allele spanned a large interval (minimum shared boundary: chr7:86,817,459-90,407,237) (**Figure 2B**) and contained 21 known genes.

**Figure 2.**
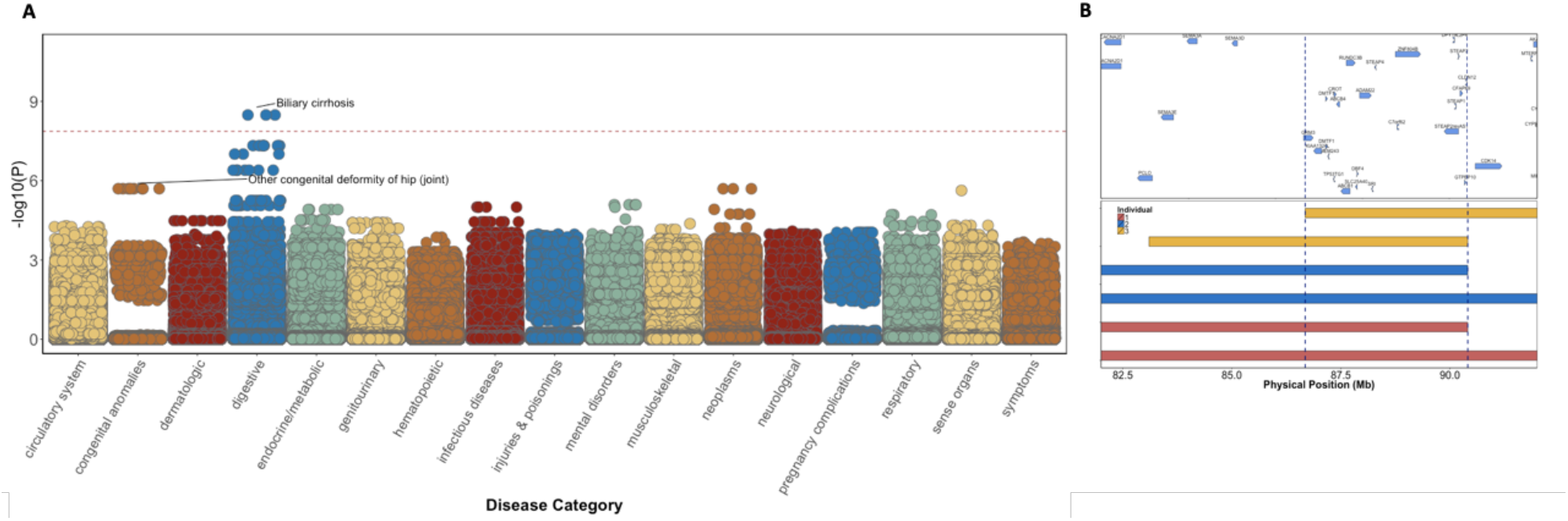
**A) Phenome-Wide Population Scale Linkage of IBD Haplotypes in Puerto Ricans.** PheWAS of recessive IBD-haplotypes against N=4880 ICD-9 billing codes revealed an association between “biliary cirrhosis” and an IBD-haplotype at 7q21.12. Red-dashed line represents the study-wide significance threshold. We also observed a suggestive association signal between an IBD-haplotype at 9q32 and “other congenital deformity of the hip”, recapitulating the known signal for Steel Syndrome in Puerto Ricans at that locus. **B)** The minimum shared boundary of the IBD-haplotypes shared between all three homozygotes at 7q21.12 spans a highly genic mapping interval of 3.6MB.

### Fine-mapping IBD-haplotypic signal uncovers a cryptic splice variant in ABCB4

To finemap the signal we performed whole genome sequencing of all three homozygous carriers, and characterized variants that fell within the minimum shared boundary of the haplotypic allele. Under the hypothesis that the causal variant would be rare we filtered to retain only variants with a global minor allele frequency of <1% (in any population group from gnoMAD or 1000 Genomes; Supplementary Table 1). We identified a total of 195 that were shared in the homozygous state between all three individuals, none of which represented non-synonymous coding variation. We found 24 sites homozygous in all three individuals that were also present in ClinVar (Supplementary Table 2). Intersecting this list with the allele frequency data, only one variant had a MAF of < 1% across all population databases, a single nucleotide variant (rs201498350, NM_000443.4(*ABCB4*):c.2784-12T>C); this variant had been asserted as “Likely Pathogenic” for “progressive familial intrahepatic cholestasis, type 1” (PFIC1) by a single submitter. The *ABCB4:c*.*2784-12T*>*C* variant has a CADD[22] score of 15.9, and a spliceAI[23] score of 0.39 (interpreted as the probability of causing a splice acceptor loss). This variant occurs in a polypyrimidine tract 12bp from the 3’ splice site of intron 22. The natural occurrence of *ABCB4* mRNAs (NM_018850.2) that lack exon 23 indicates that this splice site is weak and prone to exon skipping. This is further supported by our observation when examining HEK-293 cells tha *ABCB4* cDNA fragments are expressed both with and without exon 23 (**supplementary figure 2**). Skipping of exon 23 leads to a 141bp deletion and likely encodes for a non-functional protein due to the deletion of 47 amino acids (929 to 975), which encompasses the majority of transmembrane helix 11 and the last extracellular loop.

**Table 1.**
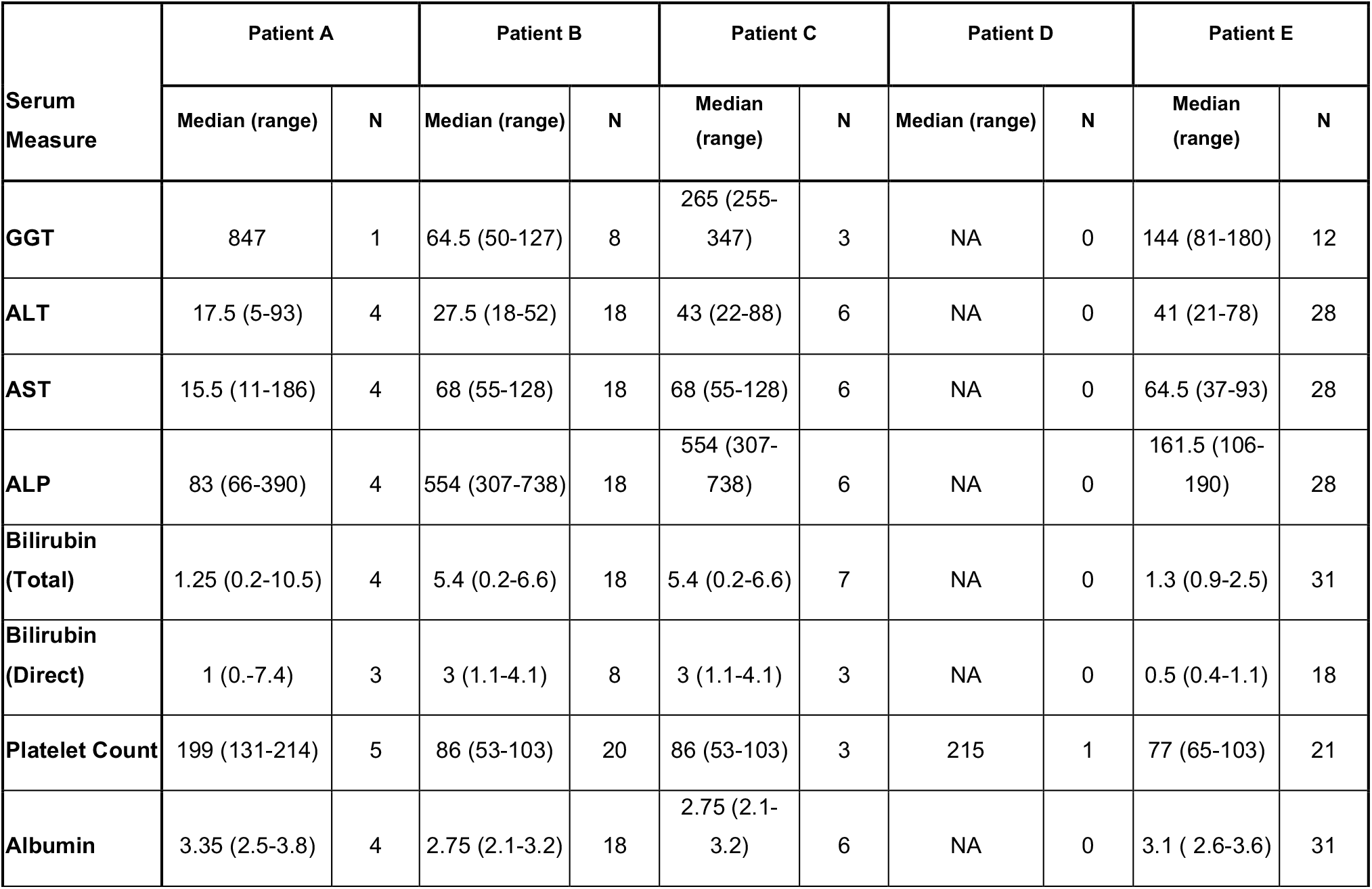
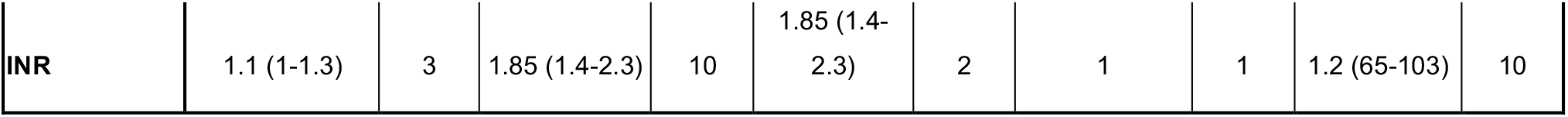
Summary of outpatient liver enzymes and liver function tests for the five *ABCB4:c*.*2784-12T*>*C* homozygotes.

**Table 2.** Phenome-wide association study of heterozygous carrier status of *ABCB4:c*.*2784-12T*>*C*.

| ICD-9 Code | Beta | Standard Error | p-value | ICD-9 Translation | Disease Category |
| --- | --- | --- | --- | --- | --- |
| 788.62 | 5.69 | 1.73 | 0.00098 | Slowing of urinary stream | genitourinary |
| 574.1 | 1.96 | 0.64 | 0.00222 | Calculus of gallbladder with other cholecystitis, without mention of obstruction | digestive |
| 724.02 | 1.66 | 0.59 | 0.00465 | Spinal stenosis of lumbar region | musculoskeletal |
| 396.3 | 5.20 | 1.85 | 0.00489 | Mitral valve insufficiency and aortic valve insufficiency | circulatory system |
| 695.9 | 2.01 | 0.72 | 0.00537 | Unspecified erythematous condition | dermatologic |
| E933.1 | 2.79 | 1.01 | 0.00547 | Antineoplastic and immunosuppressive drugs causing adverse effects in therapeutic use | injuries & poisonings |
| 640.03 | 3.34 | 1.22 | 0.00607 | Threatened abortion, antepartum | pregnancy complications |
| 680.9 | 3.43 | 1.25 | 0.00616 | Carbuncle and furuncle of unspecified site | dermatologic |
| V81.0 | 4.63 | 1.72 | 0.00694 | Screening for Ischemic Heart Disease | NA |
| 249.6 | 2.57 | 0.95 | 0.00704 | Secondary diabetes mellitus with neurological manifestations, not stated as uncontrolled, or unspecified | endocrine/metabolic |

In vivo *analysis of ABCB4:c*.*2784-12T*>*C indicates it causes increased skipping of exon 23* To test whether *ABCB4:*c.2784-12T>C affects splicing of exon 23, we cloned a genomic region of *ABCB4* containing exons 22 to 24 in an expression vector and expressed this fragment in HEK-293 cells (**Figure 3**). RT-PCR shows that the resulting pre-mRNA fragment is spliced into mRNA with and without exon 23. In this assay, the mRNA without exon 23 is more abundant than the mRNA with exon 23. Mutating the consensus T at the −12 position of intron 22 into the less-favored pyrimidine C further decreases splicing efficiency at this acceptor site. Mutating it to the purine G appears to prevent splicing completely. Our results show that the splice acceptor site of intron 22 is weak, and that the c.2784-12T>C variant increases skipping of exon 23.

**Figure 3.**
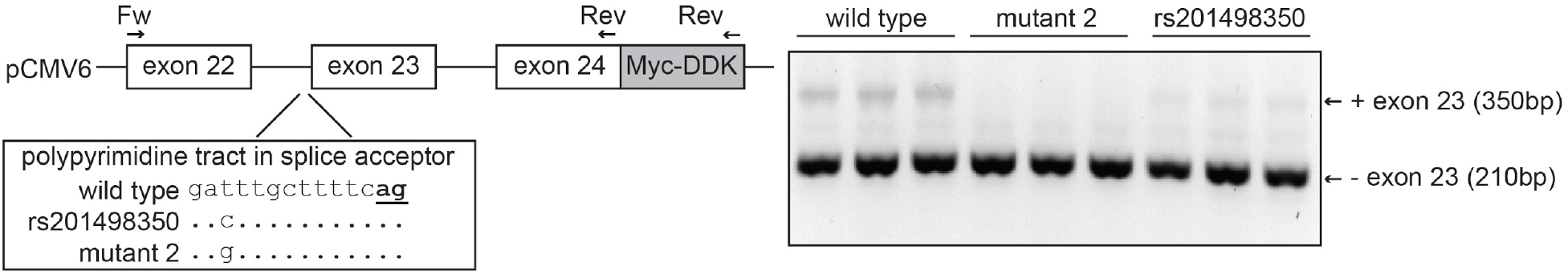
*ABCB4:*c.2784-12T>C leads to increased skipping of exon 23. Schematic representation of the approach to study the effect of *ABCB4:*c.2784-12T>C (rs201498350) on *ABCB4* splicing. A genomic region containing exons 22 to exons 24 of *ABCB4* was cloned into the pCMV6 expression vector using a forward and reverse primer as indicated. The construct harbors the 3’ 65bp of exon 22, intron 22 (1,583bp), exon 23 (141bp), intron 23 (2,500bp) and the 5’ 51bp of exon 24. The location of rs201498350 in the polypyrimidine tract in the splice acceptor site is indicated. The consensus AG at the −2 and −1 position of the splice acceptor is bold and underlined. The rs201498350 as well as another mutation (mutant 2) were introduced into the pCMV6 vector followed by transfection into HEK-293 cells (triplicate). The result of the RT-PCR with the Fw and Myc-DDK reverse primers is shown as the inverted colors of the ethidium bromide staining of a 2% agarose gel.

### Clinical characterization of ABCB4:c.2784-12T>C in homozygotes

Subsequent to the discovery of the c.2784-12T>C variant, we obtained exome sequencing data for a larger dataset of unrelated Bio*Me* participants (N=28,344). This included N=4332 PR participants who were in the original discovery dataset, and N=1015 independent PR participants. Leveraging off-target exome sequencing reads in the independent dataset, we identified two additional participants who were homozygous for the c.2784-12T>C variant. A subject domain expert performed manual chart review of all five homozygotes. Evaluation of outpatient measures of serum liver enzyme levels and liver function tests revealed significant elevation of measures consistent with liver disease (**Table 1**). Four of the five homozygotes were found to have a diagnosis of cirrhosis on chart review, and the fifth had liver steatosis on imaging. Each homozygote had a distinct etiology of their liver disease: alcohol-associated cirrhosis, primary sclerosing cholangitis, primary biliary cholangitis (with possible component of alcohol-associated liver disease) and cryptogenic cirrhosis. Two had undergone liver transplant and one was found to have an incidental hepatocellular carcinoma on explant.

### Clinical Characterization of ABCB4:c.2784-12T>C in Heterozygotes

Variation in *ABCB4* is known to confer susceptibility to hepatobiliary disease *via* both autosomal dominant (AD) and autosomal recessive (AR) modes of inheritance, and with variation in severity of disease[27],[28]. To clinically characterize the *c*.*2784-12T*>*C* variant in heterozygotes, we identified via exome sequence data N=73 PR participants in the original discovery dataset (of which N=50 were carriers of the discovery IBD haplotype which has 75% concordance with the causal variant, **Supplementary Table 3**)), and N=11 in the independent dataset of PR participants, for a total of N=84 PR heterozygotes. We compared this cohort to clinical data for N=5248 PR participants who did not harbor the *c*.*2784-12T*>*C* variant. To test for evidence of liver and other phenotypes in heterozygous carriers of *ABCB4:c*.*2784-12T*>*C*, we performed a PheWAS of ICD-9 codes. While no association achieved study-wide significance, the ICD-9 “574.10”, which encodes for “Calculus of gallbladder with other cholecystitis, without mention of obstruction” was ranked second among all associations (p<0.002; odds ratio=7.1; SE=1.9; **Table 2**). We also explored the relationship between *ABCB4:*c.2784-12T>C and nine outpatient serum liver enzyme levels and liver function tests in heterozygous carriers. We extracted these measures (**Supplementary Figure 3**) and performed linear regression of *ABCB4:*c.2784-12T>C carrier status versus the nine laboratory measures, adjusting for age and sex (**Table 3**). Both alanine aminotransferase (ALT) and aspartate transaminase (AST) were significantly elevated among carriers (p<0.0007 (beta=0.39; SE=0.21) and p<0.002 (beta=0.36; SE=0.21), respectively) after adjusting for multiple testing (study-wide significance threshold p<0.0056), while the association between *ABCB4:*c.2784-12T>C carrier status and elevated gamma-glutamyl transferase (GGT) achieved nominal significance (p<0.03). To follow up these findings, we further evaluated the association of *ABCB4:*c.2784-12T>C with liver disease phenotypes by performing manual chart review of 50 *ABCB4:*c.2784-12T>C carriers and 50 age-, sex-, and ancestry-matched non-carriers. We excluded 14 subjects with viral hepatitis from further analysis. Medical records from the remaining 43 carriers and 43 non-carriers were reviewed for evidence of any non-viral liver disease by a physician blinded to subjects’ *ABCB4* carrier status. A total of 18 of 43 carriers (41.9%) had evidence of liver disease, compared to 8 of 43 non-carriers (18.6%; p=0.03, OR=3.01). Together with the findings of advanced liver disease in homozygotes, this suggests that *ABCB4:*c.2784-12T>C is associated with increased risk of liver disease in an allele dose-dependent manner.

**Table 3.** Association study of heterozygous carrier status of *ABCB4:c*.*2784-12T*>*C* with nine outpatient serum measures.

| Phenotype | Measures | Beta | p-value |
| --- | --- | --- | --- |
| Albumin | 4309 | -0.01 | 0.956 |
| Alkaline Phosphatase | 4347 | 0.19 | 0.091 |
| ***Alanine Transaminase*** | **4367** | **0.39** | **0.0007\*\*** |
| ***Aspartate Transaminase*** | **4288** | **0.36** | **0.002\*\*** |
| Direct Bilirubin | 2875 | 0.08 | 0.566 |
| Total Bilirubin | 4331 | 0.03 | 0.781 |
| ***Gamma-Glutamyl Transferase*** | **1474** | **0.41** | **0.027\*** |
| International Normalized Ratio | 2681 | -0.03 | 0.838 |
| Blood Platelet Count | 4250 | -0.01 | 0.924 |

### Population History and Global Distribution of ABCB4:c.2784-12T>C

Finally, to gain a better understanding of which populations may harbor the *ABCB4:c*.*2784-12T*>*C* risk variant, we leveraged complete survey information on ethnicity and geographical origin. One homozygote reported being born in Puerto Rico, while the remaining four self-reported being born on the US mainland. By exploring segregation based on self-reported country of birth and self-reported ethnicity, we observed that N=42 carriers reported being born in PR (out of N=2251 PR-born individuals in total), suggesting a carrier rate of 1.95% in PR (**Figure 4A**). The remaining N=42 carriers reported being born on the US mainland, with 40 self-identifying as Hispanic/Latino (carrier rate of 1.36%). Three carriers reported being born in the Dominican Republic, one reported being born in Barbados, and the remaining two self-identifying as European American. Examination of local ancestry along the maximum shared boundary of IBD sharing between the three original homozygotes revealed all to be homozygous for European ancestry across the locus (**Figure 4B**). Additionally, the variant is present in 27 copies in the gnomAD(v3.1)[24,25] database, at a minor allele frequency of 0.16% among the “Latino/Admixed American” population, and with a single copy being present in each of the “Other”, “African/African American”, and “European (non-Finnish)” populations. We also identified a total of N=165 carriers in the UK Biobank dataset, N=163 of which self-identified as “White”, and the remaining two did not report an ethnicity in the survey. Examining carriers by country of origin, we noted that the majority self-reported being born in European countries with the highest carrier rate in Austria (0.5%) and lowest in England (0.03%) (**Table 4**). Overall this suggests that *ABCB4:c*.*2784-12T*>*C* is segregating at very low frequency in European populations, and arose to higher frequency in the PR population due to a founder effect on the European ancestral background[26].

**Table 4.** Minor allele count and carrier rate for N=165 copies of *ABCB4:c*.*2784-12T*>*C* by country of birth in the UK Biobank.

| Count | Total from country | Carrier rate | Country |
| --- | --- | --- | --- |
| 98 | 379058 | 0.026% (0.021-0.036%) | England |
| 49 | 39110 | 0.125% (0.095-0.166%) | Scotland |
| 6 | 21563 | 0.028% (0.013-0.060%) | Wales |
| 3 | 4785 | 0.063% (0.021-0.184%) | Republic of Ireland |
| 2 | 2989 | 0.067% (0.018-0.243%) | Northern Ireland |
| 1 | 196 | 0.510% (0.090-2.83%) | Austria |
| 1 | 271 | 0.369% (0.065-2.060%) | Brazil |
| 1 | 730 | 0.137% (0.024-0.771%) | Canada |
| 1 | 853 | 0.117% (0.021-0.661%) | France |
| 1 | 8 | NA | Isle of Man |
| 1 | 686 | 0.145% (0.025-0.821%) | New Zealand |
| 1 | 636 | 0.157% (0.028-0.885%) | Poland |

**Figure 4.**
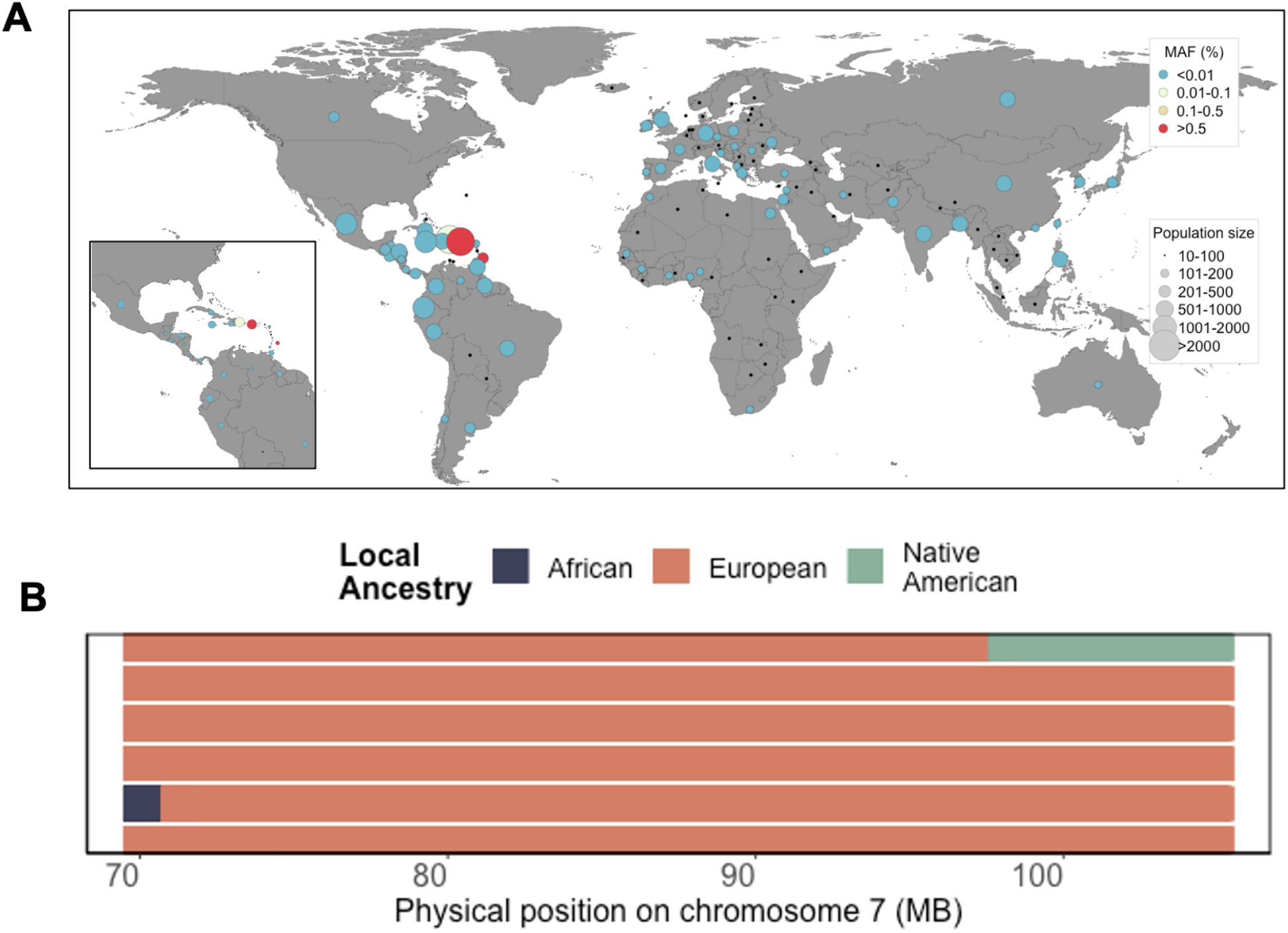
**(A) Minor Allele Frequency of *ABCB4:c*.*2784-12T*>*C* by geographic country of birth across N=134 countries in N=10232 *BioMe* participants born outside of the United States.** Population screening of *ABCB4:c*.*2784-12T*>*C* based on self-reported country of birth reveals segregation to be geographically restricted to the Caribbean, with elevated frequency in Puerto Rico. (**B**) Local ancestry across the maximum shared boundary of the three homozygotes identified through IBD sharing reveals all two be homozygous for European ancestry across the locus.

## Discussion

Here we demonstrate that by using IBD sharing to leverage distant genealogical relationships in a patient population, and by linking this to a breadth of phenotypes derived from an EHR, it is possible to discover monogenic forms of disease segregating appreciably in a large founder population. By applying this method to the Bio*Me* PR population, we uncover a genomic signal associated with liver disease. As we have previously shown, PRs represent an understudied founder population with elevated levels of cryptic relatedness, making IBD based approaches for genomic discovery especially powerful within this population.

We fine-mapped the novel signal to a non-coding variant in the *ABCB4* gene (*ABCB4:*c.2784-12T>C). We demonstrated that this variant disrupts splicing *in vitro*, resulting in an mRNA lacking exon 23 and most likely encoding a non-functional protein product. *ABCB4* encodes for the ATP binding cassette subfamily B member 4 (ABCB4), also known as multi-drug resistance protein 3 (MDR3) [29]. The protein is expressed on the canalicular membrane of hepatocytes, and is involved in the secretion of phosphatidylcholine[30], an essential component of bile, into the bile canaliculus. This role mitigates the potentially damaging effect of bile salt on the hepatocellular membrane [31,32]. Homozygous knockout mice for the murine ortholog, *Abcb4*, exhibit hepatocellular inflammation and necrosis, as well as damage to the bile ducts [31]. In humans, variation in *ABCB4* has previously been implicated in numerous forms of hepatobiliary and other liver related phenotypes [33]. Pathogenic variation in *ABCB4* is causal for progressive intrahepatic familial cholestasis type 3 (PFIC3) [34–36], a severe autosomal recessive hepatobiliary disease that typically affects children and adolescents [28]. Notably, *ABCB4* has also previously been implicated in cryptogenic cirrhosis of the liver [37]-[38], as well as a range of milder phenotypes including intrahepatic cholestasis of pregnancy [39],[40–42], drug induced liver injury[43], and low phospholipid-associated cholelithiasis[44]. The range of Mendelian *ABCB4* associated phenotypes has been noted to follow both autosomal dominant and recessive modes of inheritance. Furthermore, in large-scale population-based studies, variation in *ABCB4* has been statistically associated with elevated risk for a number of liver-related phenotypes, including risk for non-alcoholic fatty liver disease[45], elevated serum liver enzyme levels[46,47], and risk for hepatobiliary carcinoma[48], suggesting that common variation in this gene may play a broader role in liver disease risk in the human population.

A previous clinical study of PR ancestry patients with PFIC noted a high prevalence of symptoms representative of ABCB4 deficiency, and suggested an *ABCB4* founder variant may contribute to the prevalence of PFIC in the PR population [49]. We observe that *ABCB4:*c.2784-12T>C segregates at a carrier rate of 1.95% in PR ancestry individuals, while being rare or absent in non-Caribbean populations, highlighting the importance of including individuals of diverse ancestry in genomic research. The high carrier rate in PR suggests the existence of hundreds of homozygous individuals who may be at elevated risk for liver disease. We identified five homozygotes for *ABCB4:*c.2784-12T>C in the Mount Sinai health system, and found evidence of liver cirrhosis in four. The etiology of liver disease in each was noted to be different, which suggests that *ABCB4:*c.2784-12T>C could predispose to various forms of liver disease. We also noted significant elevation of liver enzyme levels in heterozygotes, as well as increased rates of liver diseases in heterozygotes compared to matched non-carriers. This suggests that heterozygous carriers of this variant are also at risk for liver disease. This is consistent with the known AD and AR inheritance of other *ABCB4* variants. Taken together, this work demonstrates the utility of genetics-first approaches to discovery in health systems for uncovering a continuum of genomic risk for common diseases. Understanding such genetic risk factors at an individual level will be useful in clinical risk stratification and care in the future.

## Methods

### BioMe Biobank

Study participants were recruited from the Bio*Me* Biobank Program of The Charles Bronfman Institute for Personalized Medicine at Mount Sinai Medical Center from 2007 onward. The Bio*Me* Biobank Program (Institutional Review Board 07–0529) operates under a Mount Sinai Institutional Review Board-approved research protocol. All study participants provided written informed consent.

### Genotype Data and Quality Control

Genotyping, quality control and merging of array data across the OMNI and MEGA platforms was performed as described in detail in Vishnu *et al*. 2019 [50]. In brief, we performed standard quality control for variants based on missingness, heterozygosity, and Hardy Weinberg equilibrium using PLINKv1.9 [51,52]. We removed samples that were duplicated across both arrays and subset data to the intersect of variants present on both platforms (n=461,677 SNPs; n=21,692 individuals). After subsequently removing palindromic sites with a missingness rate of >1%, this resulted in a total of 377,799 SNPs and 25,750 individuals for downstream analysis.

### Haplotype Phasing

Phasing was performed per chromosome with the EAGLEv2.0.5[53] software using the genetic map (hg19) that is included in the EAGLEv2.0.5 package (url: https://data.broadinstitute.org/alkesgroup/Eagle/downloads/Eagle_v2.0.tar.gz). An additional 2 individuals were excluded during the phasing process if they had a per chromosome level missingness rate of greater than 10% for any one autosome, leaving N=25,748 individuals in total.

### Identity-by-Descent Inference and Quality Control

Phased output from EAGLE was filtered to a MAF of >= 1% and converted to PLINK format using fcGENE [54]. This was used as input for the GERMLINE algorithm [55]. We ran GERMLINE over each autosome across all individuals simultaneously using the following flags: *“-min_m 3 - err_hom 0 -err_het 2 -bits 25 –haploid*.” For quality control, IBD that overlapped with low complexity regions were excluded, along with IBD that fell within regions of excessive IBD sharing (which we defined as regions of the genome where the level of pairwise IBD sharing exceeded 3 standard deviations above the genome-wide mean).

### Identity-by-descent based clustering of Puerto Rican ancestry participants

We summed IBD haplotypes along the genome of all N=25748 participants and used to construct an adjacency matrix where each node represented a Bio*Me* participant and each weighted edge represented the pairwise sum of IBD sharing between a given pair of individuals. After first excluding edges sharing >=1500cM of their genome IBD, we employed the *InfoMap[56,57]* as implemented in the iGraph package (R version 3.2.0) to uncover communities of individuals enriched for IBD sharing. We uncovered a community of N=5100 individuals who, based on self-reporting labels, we defined as the Puerto Rican ancestry IBD-community going forward.

### Phenome Wide Association of Identity-by-Descent Haplotypes

We first clustered IBD haplotypes inferred via GERMLINE into homologous cliques using the DASH [58] *advanced (dash_adv)* algorithm across all Bio*Me* participants, including the following additional parameters: “*-win 250000 -r2 1”*. We then extracted the Puerto Rican community (n=5100) from the DASH output and recoded individuals who were homozygous for a given IBD clique as “1” and those who were heterozygous or who were not members of the clique as “0”. We then used this as the primary predictor variable for an IBD-based Phenome Wide Association that was modeled using an implementation of the Saddle Point approximation[59] (using the R package “SPAtest”, R version 3.2.0), with age and sex included as covariates. For each test, one individual from each pair of directly related individuals was excluded prior to association, preferentially excluding ‘controls’ to ‘cases’ for each ICD-9 code.

### Whole Genome Sequencing, Variant Calling and Annotation of IBD Homozygotes

Alignment and variant calling of Whole Genome Sequence (WGS) data was performed using the pipeline provided by Linderman *et al*.*[60]*. Further variant annotation was performed using Variant Effect Predictor. These annotations were then intersected the WGS data for the three homozygotes using an in-house python script.

### Phenome Wide Association of ABCB4:c.2784-12T>C in heterozygous carriers

A Phenome-wide association of *ABCB4:*c.2784-12T>C carrier status was conducted using the SAIGE software[50,61] for a total of n=4903 Puerto Rican ancestry participants (homozygous individuals were excluded). ICD-9 billing codes served as the phenotypic outcome, and we included age, sex and the first five principal components (PCs) as covariates, as well as a General Relatedness Matrix (GRM) to account for relatedness. The association analysis was restricted to ICD-9 codes for which 3 or greater cases were present among carriers (N=550 ICD-9 codes).

### Association of ABCB4:c.2784-12T>C and Liver Enzymes

Outpatient values for nine laboratory tests for liver enzymes and liver function were extracted from EHRs. For each individual, the median value was taken for each trait. Patients were stratified according to sex, and outliers that fell greater than 4 standard deviations from the sex-specific population median were excluded. Sex-specific values were subsequently log-transformed, and converted to z-scores (mean 0, standard deviation 1) before the data was recombined. These z-scores were then used as the phenotypic outcome in a linear model that included age as a covariate. Related individuals were excluded from the analysis, as were the five individuals who were homozygous for the *ABCB4:*c.2784-12T>C variant.

### Association of ABCB4:c.2784-12T>C and Liver Disease

Manual chart review was performed by a physician blinded to the subject’s ABCB4 carrier status. Subjects with hepatitis C causing viral hepatitis were excluded from further analyses. Text search was done for “liver disease”, “fatty liver”, “NAFLD”, “fibrosis”, “steatosis”, “sclerosing cholangitis”, and “cirrhosis”. A review of all prior abdominal imaging was performed, specifically assessing for phrases such as “nodular” or “hyperechogenic’’ liver. If any of these searches yielded a positive result, then clinical notes, alcohol history, BMI, liver function tests, FibroScan results, and any liver biopsies were reviewed to establish the etiology and severity of the subject’s liver disease. A two-tailed Fisher’s exact test was performed to assess for associations between carrier status and the presence of any non-viral liver disease, and a P-value of < 0.05 was considered significant.

### Functional validation of ABCB4:c.2784-12T>C

We amplified (PrimeSTAR GXL DNA Polymerase, Takara Bio) and cloned a 4,340bp *ABCB4* genomic region from exons 22 to exons 24 into the pCR2.1-TOPO vector (TOPO TA cloning kit, Invitrogen) using the following forward and reverse primers: 5’-**<u>GCGATCGC</u>**C ATG GTG TCT TTG ACC CAG GAA AGA AA-3’ and 5’-**<u>ACG CGT</u>** AGA ACT GGC ATG TCC TAG AGC C-3’. Sequence verified pCR2.1-TOPO with this fragment was used as a template to re-amplify the insert (PrimeSTAR GXL DNA Polymerase, Takara Bio) using the following forward and reverse primers: 5’-CAC TTG **<u>GCG ATC GC</u>**C ATG GTG TCT TTG ACC CAG GAA AGA A-3’ and 5’-GAT AAC **<u>ACG CGT</u>** AGA ACT GGC ATG TCC TAG AGC C-3’. The primers introduce a 5’ AsiSI/SgfI and 3’ MluI restriction site (bold and underlined) that were used for cloning the fragment into the pCMV6-entry vector (Origene). The c.2784-12T>C variant was introduced using site-directed mutagenesis (Q5 Site-Directed Mutagenesis kit, NEB) with the following oligonucleotides primers: Q5-Fw 5’-AGTATACTGAcTTGCTTTTCAG-3’ (mutated nucleotide in lower case) and Q5-Rev 5’-TGTAACCATCTCTTCAGC-3’. The wild type and variant pCMV6-ABCB4 were sequenced to confirm the absence and presence of the variant. Both vectors were transfected into HEK-293 cells using Lipofectamine 2000. After 24 hours, cells were lysed in QIAzol and RNA isolated (RNeasy mini kit, QIAGEN). RNA was used for cDNA synthesis (SuperScript IV First-strand Synthesis System, Invitrogen) after which the splicing of exons 22-24 was studied using PCR. Because HEK-293 cells express low levels of native *ABCB4*, we used the forward primer annealing in exon 22 used for cloning and a reverse primer on the MYC-DDK tag of the pCMV6 vector: DDK reverse 5’-CCT TAT CGT CGT CAT CCT TGT AAT CC-3’. All PCR fragments were Sanger sequenced to confirm their identity.

## Data Availability

The exome sequencing datasets generated and/or analyzed during the current study are not publicly available, but summary statistics are available from the corresponding author on reasonable request.

## Main Figures

**Supplementary Figure S1.**
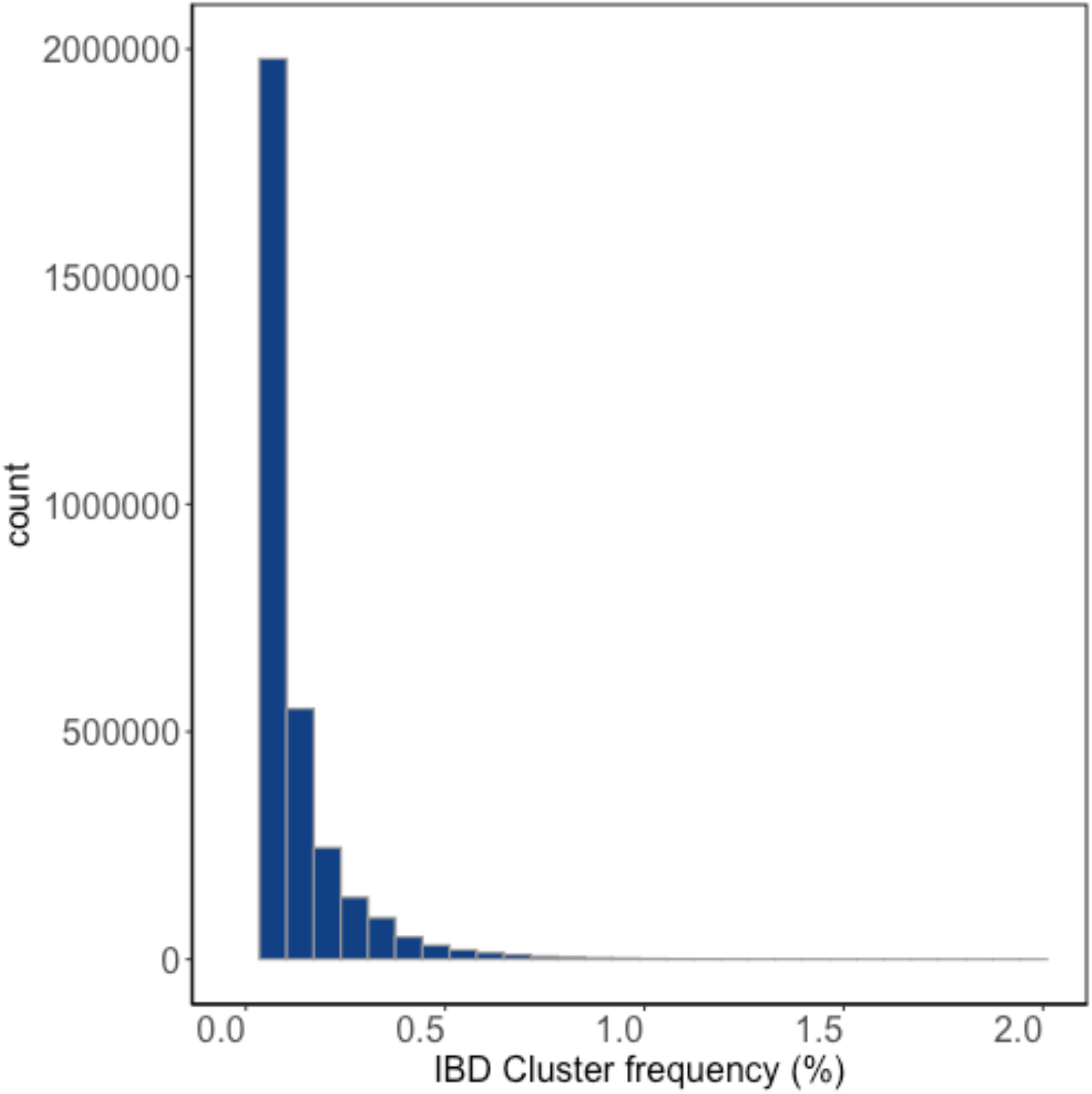
Site frequency spectrum of homologous IBD clusters in Bio*Me* PR.

**Supplementary Figure S2.**
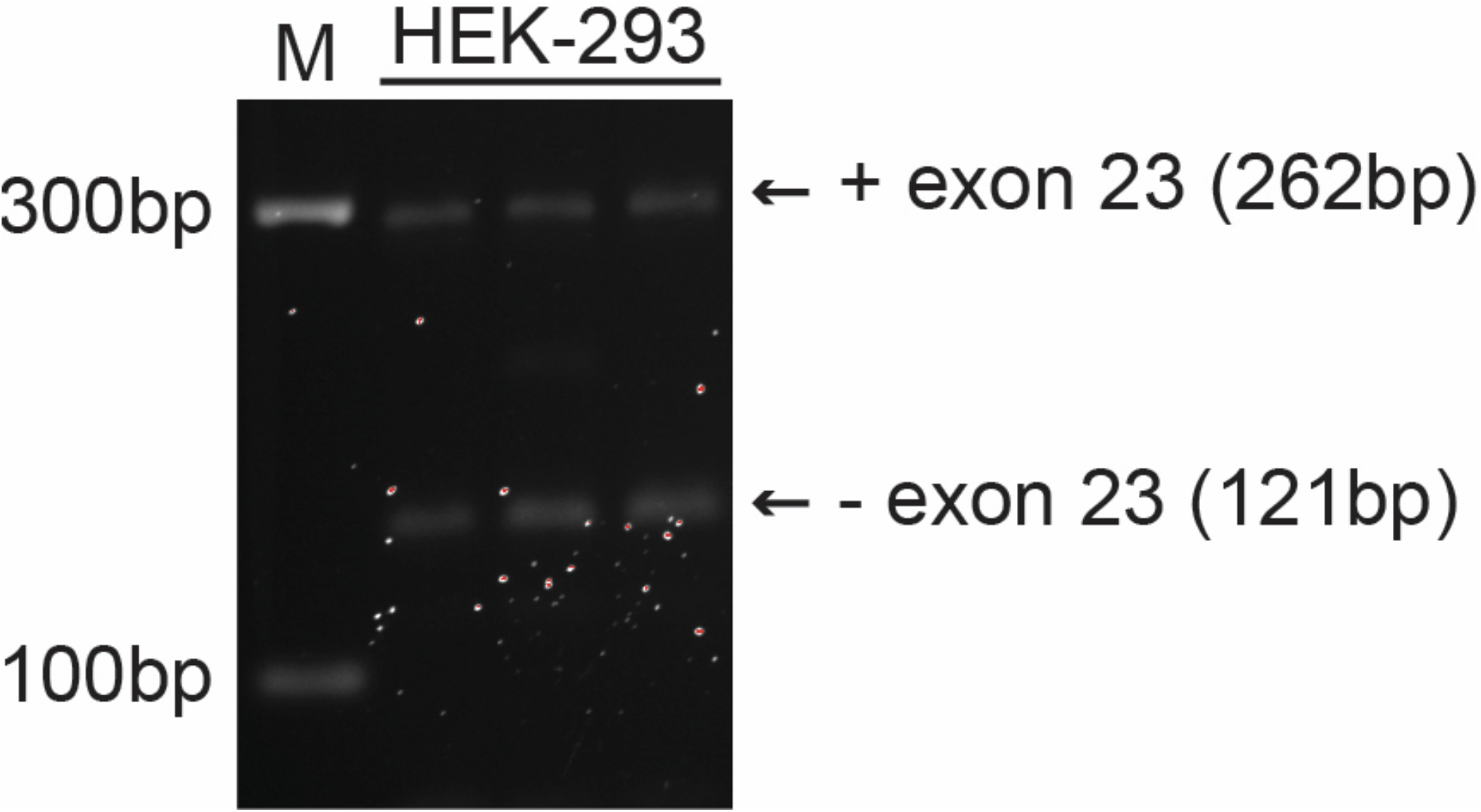
Natural occurrence of *ABCB4* cDNA lacking exon 23 in HEK-293 cells. *ABCB4* cDNA fragments were amplified from HEK-293 cDNA using primers targeting exon 22 and exon 24. The first lane contains the molecular-weight size marker. The size of the cDNA fragments are indicated. The cDNA fragments run at a slightly larger molecular-weight because the primers were tagged with additional nucleotides for cloning purposes.

**Supplementary Figure 3.**
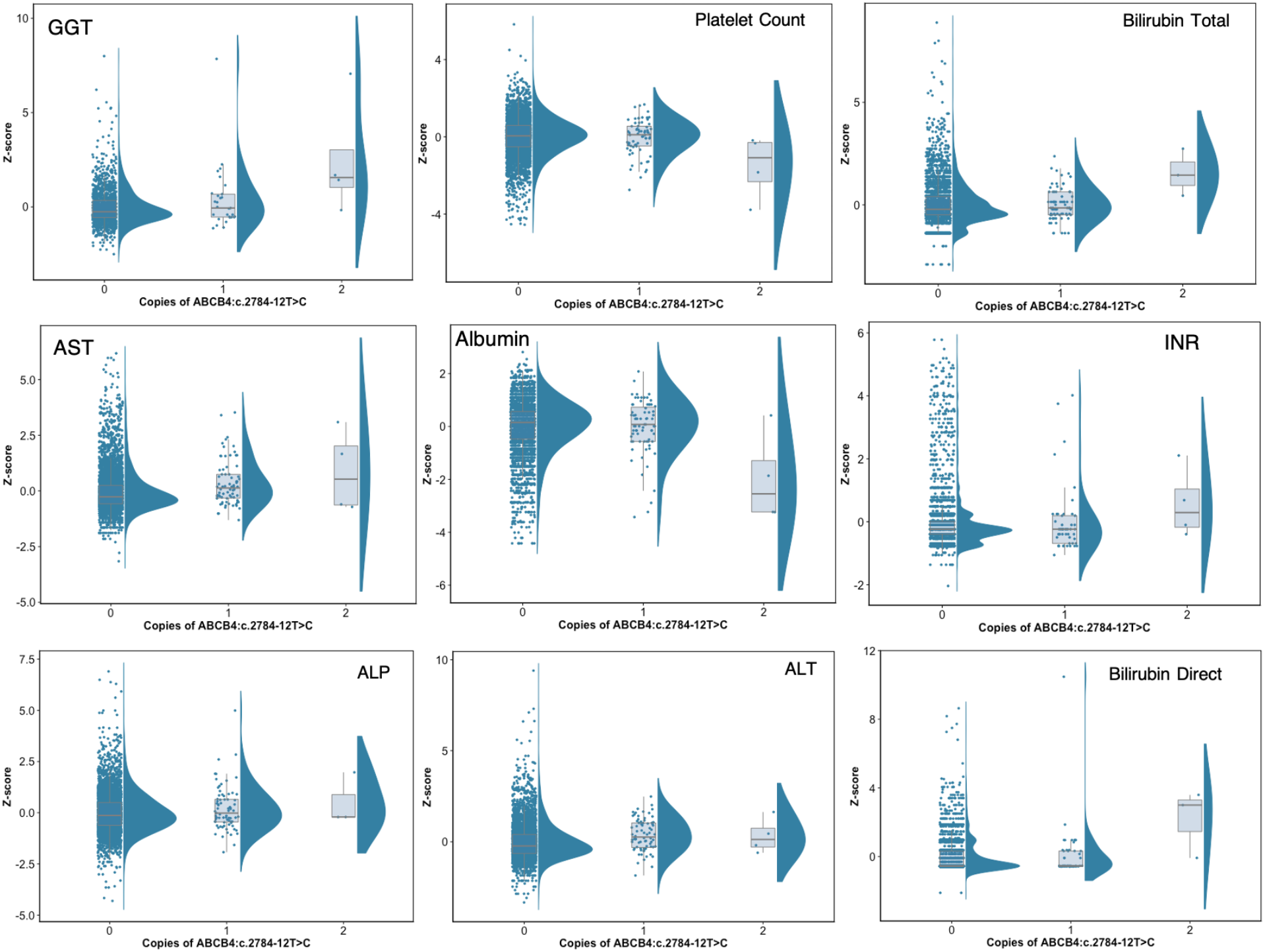
**Distributions of Z-scores for outpatient values for nine serum measures in Puerto Rican ancestry Bio*Me* participants, stratified by *ABCB4:c*.*2784-12T*>*C* carrier status.**

**Supplementary Table 3.** **Concordance between being heterozygous for the discovery IBD-haplotype and *ABCB4:c*.*2784-12T*>*C* carrier status among PR ancestry individuals present in both the genotype and exome sequence data.**

|  | IBD Carrier | IBD Non-Carrier |
| --- | --- | --- |
| **Exome Carrier** | **50** | **23** |
| **Exome Non-Carrier** | **11** | **4248** |

